# Geographical targeting of active case finding for tuberculosis in Pakistan using artificial intelligence software (SPOT-TB): a pragmatic stepped wedge cluster randomized control trial

**DOI:** 10.64898/2026.05.20.26348577

**Authors:** Amna Mahfooz, Abdullah Latif, Syed Mohammad Asad Zaidi, Wasim Ahmed, Nainan Nawaz, Tahira Ezra Reza, Adeel Tahir, Fazal ur Rehman, Shahid Naveed, Alina Shahid, Frhan Ali, Faran Emmanuel

**Affiliations:** Center for Global Public Health, Pakistan; Mercy Corps, Pakistan; UCL Centre for Global Tuberculosis Research and WHO Collaborating Centre for Tuberculosis Research and Innovation, Institute for Global Health, University College London, London, United Kingdom; University of Manitoba, Canada

## Abstract

**Background:** Community-wide active case-finding (ACF) is being increasingly implemented as a tuberculosis (TB) elimination intervention. However, conventional site selection strategies may result in low yields from screening. We evaluated whether an artificial intelligence (AI) software guided targeting strategy could improve detection of TB during screening activities (called camps) relative to routine approaches to site selection in the programmatic setting in Pakistan.

**Methods:** We conducted a stepped-wedge cluster-randomised trial embedded within Global Fund supported ACF activities implemented by Pakistan’s National TB Program and private sector partners. Thirty mobile X-ray van teams operating in 68 districts were randomly assigned to transition from routine site selection approaches (based on field-staff experience and historical data) to an AI-guided targeting strategy, using the software MATCH-AI. We assessed the effect of the intervention on the primary outcome, Camp Positivity Yield, defined as the number of individuals diagnosed with bacteriologically confirmed TB per camp, using generalised linear mixed models. The primary analysis was by intention to treat. Camps conducted within a 5-km radius of the AI selected locations were included in a “validated” per-protocol analysis. We conducted several district-level subgroup analyses. This trial is registered, number NCT06017843.

**Findings:** Between August 2023 and September 2024, 3,936 screening camps were conducted (2,046 control, 1,890 intervention), screening 269,254 individuals. In the intention-to-treat analysis, Camp Positivity Yield was 7% higher in the intervention group relative to the control group, however this difference was not statistically significant (adjusted risk ratio [RR] 1·07, 95% CI: 0·94–1·22). In the validated per-protocol analysis, Camp Positivity Yield was 32% higher in the intervention group relative to the control group (adjusted RR 1·32, 95% CI: 1·12–1·54). Yields were highest in districts that had moderate baseline yields of 0.5-1% per population screened prior to the trial (adjusted RR: 1.57, 95% CI: 1.13 – 2.18) and in rural districts (adjusted RR 1.43, 95% CI: 1.23 – 1.65).

**Interpretation:** The use of an AI-guided targeting strategy significantly increased detection of bacteriologically confirmed TB during active case-finding in the validated per-protocol analysis, relative to conventional site-selection approaches employed by field-staff. This software may be considered as a supportive tool to improve the efficiency of community-based TB case-finding interventions in other high burden countries.

**Funding:** This study was funded by the Bill & Melinda Gates Foundation grant number INV-037454. The study was embedded within ongoing active case finding (ACF) activities supported operationally by the Global Fund through the National TB Control Program Pakistan.

**Research in context:** *Evidence before this study:* We searched PubMed for articles published between Jan 1, 2010, and Dec 31, 2024, using the terms “tuberculosis”, “surveillance”, “active case finding”, “community-based screening”, “hotspot targeting”, “predictive modelling”, and “artificial intelligence”. A previously conducted systematic review by Cudhay *et al*. in 2018 identified three prospective studies using spatially targeted interventions for TB from low incidence settings but none from high incidence settings. One study reported from Uganda in 2025 identified a moderate benefit of conducting door-to-door screening in a 50-100m radius around households of tuberculosis (TB) cases diagnosed from passive case-finding in health facilities, relative to the general population. We identified several retrospective analyses of screening interventions identifying spatial variation in TB case-detection. However, no prospective studies or randomized evaluations were identified that compared methods for selecting geographic areas for screening, including through data-driven or artificial intelligence (AI) based approaches.

*Added value of this study:* This stepped-wedge cluster randomized trial is the first prospective evaluation of a geographical targeting strategy for TB screening from a high incidence setting. The trial compared screening in hotspots predicted by an artificial intelligence software (MATCH-AI) to the standard of care that largely relied on staff-led approaches and historical experience for site-selection in Pakistan. We found that use of AI software increased detection of bacteriologically confirmed TB relative to conventional site selection methods in the “validated” per-protocol analysis in which screening locations were verified independently to be within a 5-km radius of hotspots identified by the software. These findings provide further evidence for spatial variation in TB burden and support the strategy of targeted screening in geographic hotspots. This trial was conducted on a national scale and under real-world programmatic conditions. Our results provide empirical evidence in support of a commercially available software that programs in other high-burden, low-resource settings can utilize to improve the efficiency of community-based screening through systematic and data-driven approaches.

*Implications of all the available evidence:* Globally between 1.9 – 3.5 million incident TB cases are not diagnosed each year resulting in significant morbidity and mortality that may be preventable. Early diagnosis and treatment of these individuals may also prevent community transmission and support TB elimination. There is significant evidence that distribution of TB is spatially heterogeneous, however robust, quantitative approaches towards identification of hotspots using routine surveillance data had been challenging. Our study demonstrates that AI can enhance the efficiency of active case-finding interventions within routine programmatic activities without requiring expansion of screening coverage. For programs operating under resource constraints such as in Pakistan, integrating such tools into routine operational planning may allow screening activities to be more strategically deployed to populations at highest risk. This may improve early diagnosis and treatment of individuals with TB and help accelerate TB elimination efforts in high burden countries with substantial gaps in case detection.

## Introduction

In 2024, an estimated 10·7 million people fell ill with tuberculosis (TB) and 1·23 million people died from the disease ^1^. A significant proportion of people with TB remain undiagnosed and untreated, sustaining transmission and preventing disease elimination ^1^. In the same year, Pakistan had a 25% gap between estimated incident and notified cases and the country contributed 6·3% to the global TB burden ^1,2^. Community-wide screening interventions, such as active case finding (ACF) can support facility-based services (passive case finding) in case-detection and may help reduce TB incidence and prevalence through potential detection of early and asymptomatic TB ^3–8^.

Following national policy shifts to expand active case finding using mobile chest X-ray, Pakistan has increased efforts to close the case-detection gap by conducting ACF at scale through screening events called “chest camps” or simply “camps” using mobile vans fitted with digital chest X-rays (CXR). Screening is supported by computer-aided detection (CAD) software and confirmatory diagnosis is conducted using molecular testing through GeneXpert machines within vans or at fixed facilities ^4,9,10^. Between 2019 and 2024, a total of 12,264 screening camps were conducted across 96 districts, supported by an expanding fleet of mobile X-ray vans, which grew from 3 in 2016 to 36 in 2024. The programme scaled rapidly over time, with 4,632 camps conducted in 2024 alone. Across this period, 564,862 individuals underwent chest X-ray screening, representing 72.2% of all participants attending these camps. However, as screening activities are expanded to cover the general population, yields from ACF tend to decrease ^11^. There is evidence to suggest that the effectiveness of these camps may be diluted by suboptimal geographic targeting and previous studies from Pakistan have highlighted significant spatial variation in TB risk at the regional and neighborhood levels^12,13^. The 2019 TB Joint Programme Review Mission (JPRM), conducted by the Government of Pakistan together with the World Health Organization (WHO) and other partners, highlighted lower than expected yields from ACF and recommended adopting a more geographically targeted approach in areas where TB prevalence exceeds WHO recommended thresholds ^14^.

Responding to these recommendations, the National TB Control Program (NTP) implemented the use of MATCH-AI (Mapping and Analysis for Tailored disease Control and Health system strengthening-Artificial-Intelligence), a decision-support software tool that integrates programmatic data and contextual variables to predict areas of high TB risk and potential hotspots through Bayesian modeling ^15,16^. MATCH-AI was launched in mid-2019 and implemented programmatically in selected districts of Pakistan through routine active case finding activities led by the National TB Control Program in partnership with Mercy Corps (the private-sector implementing partner of NTP), but like all such decision support tools for targeting screening activities, it lacked evidence for efficacy and cost-effectiveness. Deployment of such tools has important programmatic implications in both cost and operation. We therefore sought to evaluate whether the use of an AI-guided geographical targeting tool to plan screening camp deployment increased detection of bacteriologically confirmed TB compared with conventional site selection methods.

## Methods

### Study design and participants

We implemented a stepped-wedge cluster randomized trial in 68 districts and four provinces of Pakistan using a fleet of 30 mobile X-ray vans and associated field-teams (Figure 1). The trial was embedded within programmatic Global Fund supported community ACF activities implemented by NTP in partnership with Mercy Corps and sub-recipient organizations.

**Figure 1:**
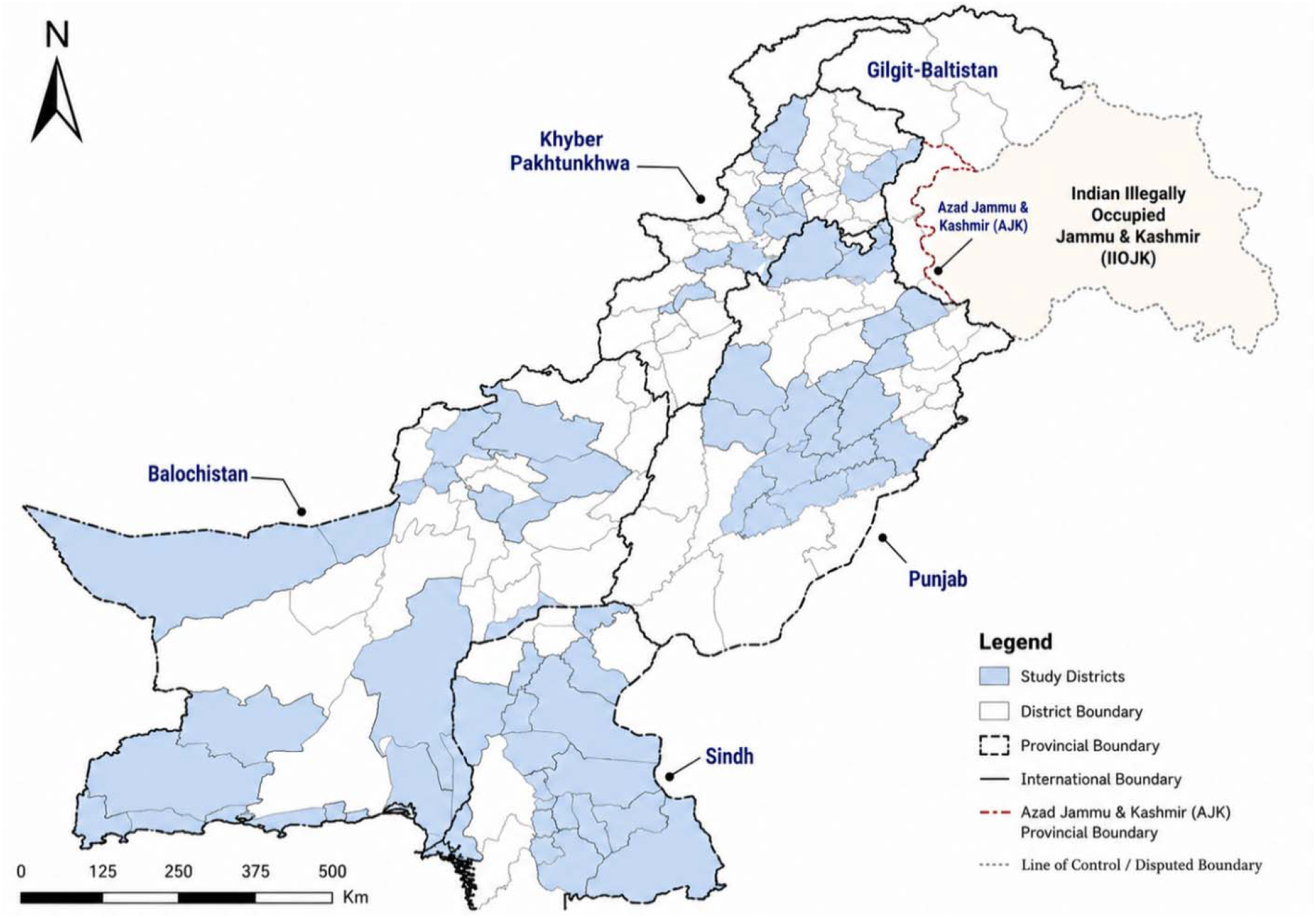
Study Districts.

The study population comprised all individuals attending community ACF camps organized by the field-teams during the study period. Screening procedures have been previously described in detail ^17^. Eligibility for screening was regardless of symptoms. Children (<12 years of age) and pregnant women were excluded from chest X-ray screening as per the national guidelines but were eligible for symptom screening and further diagnostic evaluation if symptomatic. Individuals ineligible for chest X-ray screening contributed to camp attendance counts and downstream diagnostic outcomes where applicable but were excluded from CXR-based denominators. All screening, diagnostic testing and case notifications procedures followed existing NTP guidelines and were unaffected by this trial. Mobilization activities such as distribution of promotional material, announcements, meetings with community-notables, such as, village elders, local politicians, imams of mosques, etc., were unaffected by this trial.

### Procedures

In the control arm, field teams continued to use existing approaches for camp site selection. Every month, the ACF department at Mercy Corps assigns targets for the number of screening camps to be conducted in each district to implementing partner organizations who are Global Fund sub-recipient (SRs). While SRs are expected to meet these targets, the selection of camp locations and timing is not formally prescribed by the National TB Control Program or Mercy Corps. Instead, site selection is guided by local stakeholder consultation and led by District Field Supervisors in coordination with District TB Coordinators, drawing on local knowledge, TB notification data, previous camp performance, and historical paper-based TB registries. Camps are typically conducted in underserved urban and rural areas, and are often organized in collaboration with local non-profits and private healthcare facilities such as general practitioner (GP) clinics. However, site selection approaches were not systematic and did not use consistent quantitative methods to identify TB hotspots, with practices varying across SRs based on local context and operational feasibility.

In the intervention group, camps were conducted primarily in locations guided by MATCH-AI. Details on the prediction model used for MATCH-AI have been described elsewhere ^1^. Briefly, the software divided the country into 10,000 population polygons and predicted the risk of TB based on a range of contextual variables (such as income, population density, immunization coverage, etc.) and programmatic surveillance data. The software interface allows for visualization of high-risk areas and provides GPS locations and street addresses for the centroids of each polygon. Field-teams were provided with a list of the highest predicted TB prevalence polygons in each district as predicted by MATCH-AI and instructed to conduct camps at those locations. Teams were provided up to 15 locations per district, and these remained unchanged for the duration of the trial. The teams were asked to choose the highest ranked recommended locations for screening; if for some reason they were unable to pick that location, they were instructed to choose the next one. Once van-teams had switched over to the intervention group, they continued to use MATCH-AI to guide their decision-making through to the end of the trial. A minimum (unspecified) number of camps were allowed independent of the MATCH-AI predictions due to any prior commitments of van-teams with local communities or due to logistical constraints in reaching AI-directed sites.

### Randomization and masking

During the initial baseline period for two months (August–September 2023), all 30 vans operated in the control arm. From October 2023, three vans were randomly assigned each month to transition into the intervention arm, where site selection was guided by MATCH-AI recommendations. The trial data manager generated a computerized randomization sequence before the start of the trial. The trial experienced a pause (due to programmatic issues) between December 2023 and January 2024, and the activities resumed in February 2024. By September 2024, all 30 vans had transitioned to the intervention arm.

Since field-teams were directed to the screening sites, they could not be masked to the intervention status. However, they were masked to the randomization sequence and the timing of their van’s transition from control to intervention until 15 days before implementation. Individuals attending screening camps were not informed of the intervention status of the site, however, they were informed that screening data may be used for research purposes during consent for chest X-ray screening.

### Data sources and validation

We combined data from three primary sources to generate the final dataset for analysis. First, implementer datasets in the form of programmatic reports at the conclusion of each camp were aggregated. Nine sub-recipient partners of Mercy Corps submitted periodic datasets (monthly, quarterly, or biannually) describing camp activities including the number of participants, X-rays performed, presumptive identified, Xpert tests done, and diagnostic outcomes using paper-based forms. These datasets varied in format and completeness, requiring manual harmonization and variable mapping. For the intervention camps, site recommendations from the MATCH-AI software were mapped on to individual vans and standardized to produce a master recommendations dataset comprising of approximately 6,761 recommended sites for the 30 van teams. Finally, these were matched to actual locations of camp sites and validated using WhatsApp logs sent by field-teams from the camp sites. WhatsApp messages for each camp contained camp and van IDs, camp address and a GPS coordinate link using Google Maps. These were used to generate a structured location dataset of AI-guided camps (1,095 camp locations). Locations shared by field-teams were used to calculate the distances of the actual camp sites from the MATCH-AI recommendations and determined which camps were within a 5-km radius. These three data sources allowed us to monitor intervention fidelity and provide feedback to field-teams. Because data streams were decentralized and heterogeneous, we applied consistent cleaning rules before merging into the master analytic dataset.

### Sample size

Since this intervention was being rolled out programmatically, the sample size was not capped and all camps in the study period were included from vans meeting the eligibility criteria. Using camp data from 2022, we calculated a mean of 0·88 bacteriologically confirmed TB (B+) cases diagnosed⍰per camp with an over dispersion of 1·34 and an intra-cluster correlation (ICC) of 0·12. We used a cross-sectional sampling structure, assuming an exchangeable correlation and a significance level of 0·05. Based on retrospective analysis of camps data, we anticipated that the 30 vans would conduct 3,300 camps during the study period. This sample size provided a power of 80% to detect a 20% difference in Camp Positivity Yields between the intervention and control arms. These calculations were intended to provide an approximate indication of power in a pragmatic stepped-wedge design rather than a fixed target for the sample size.

### Outcomes

The primary outcome was Camp Positivity Yield, defined as counts of bacteriologically confirmed TB (B+) cases diagnosed in each camp. Secondary outcomes included Camp Positivity Rate defined as B+ cases diagnosed per population screened with CXR, Camp All-Forms Yield defined as counts of All-Forms TB (AF-TB) cases diagnosed in each camp and camp All-Forms TB Rate defined as AF-TB cases diagnosed per population screened with CXR. Due to lack of standardization in clinical practice in Pakistan, B+ TB was considered a more reliable measure, relative to AF-TB that included individuals with bacteriologically unconfirmed TB as well as extra-pulmonary TB who are treated empirically by clinicians. Yield per camp rather than per population screened was chosen for the primary outcome as this metric directly reflects the operational efficiency of the ACF intervention. Microbiological outcomes were based on Xpert testing results conducted in the mobile X-ray vans or at fixed public-sector TB laboratories.

## Statistical analysis

We used frequency statistics to describe and compare the baseline characteristics of camps and participants, as well as the screening care cascade, between the intervention and control groups. We utilized t-tests to compare means for continuous variables and Pearson’s chi-squared tests for categorical variables. A generalized linear mixed negative binomial regression was utilized to model the counts of B+ TB diagnosed per camp (due to over-dispersion of data) and a generalized linear mixed Poisson regression was utilized to model counts of AF-TB. To account for secular trends, a fixed effect for time was added into the model as a categorical variable. To account for clustering of outcomes within the randomization units, the van teams were added as a random effect in the model. Both adjustments were pre-specified and have been described further in the protocol statistical analysis plan. Community notables meeting was added as a dichotomous explanatory variable to adjust for pre-camp community mobilization activities. To calculate the B+ and AF-TB Rate outcomes, the log of X-rays conducted in each camp were used as an offset in the regression models. Camps were categorized into intervention or control based on their exposure to the MATCH-AI intervention and added as a binary explanatory variable into the model. The model calculates incidence rate ratios between the two arms to determine the intervention effect and these were presented as “Risk Ratios” and “Rate Ratios” where appropriate, with 95% Confidence Intervals (CI).

The primary analysis was intention-to-treat (ITT) where all camps were included once teams had switched to the intervention group after their assignment, regardless of whether those camps were conducted using MATCH-AI. The per-protocol (PP) analysis included intervention camps that were reported by the van teams (using camp reports) as having been conducted using MATCH-AI– recommended locations. However, since self-reported adherence may not have fully reflected whether teams implemented camps at the recommended sites, we conducted a “validated” per-protocol analysis. In this stricter analysis, intervention camps were included only if their GPS coordinates confirmed that the camp was conducted within a 5-km radius of the specific location recommended by MATCH-AI, as verified independently by the study team. The primary analysis was ITT; validated per-protocol results are presented alongside ITT estimates in the main manuscript, with additional analyses in the appendix. The per-protocol analysis results have not been presented. Camps in the control arm remained the same across all analyses.

A pre-specified subgroup analysis was carried out by stratifying the effect of the intervention over district-level yields of TB (high, medium, low) identified through a retrospective analysis of mobile X-ray-based camps in 2022 (using which the sample size calculations were performed) and a recently published district-level analysis of ACF yields in Pakistan. Districts with yields below 0·5% were classified as “Low-Yield”, those with 0·5% - 1% as “Medium-Yield” and those with >1% as “High-Yield.” Districts where screening data was not available were classified as Medium Yield. All post-hoc analyses were considered exploratory and were conducted for the validated per-protocol camps by each of Pakistan’s four major provinces (Punjab, Sindh, Khyber-Pakhtunkhwa and Balochistan) and by district population density, with districts categorized as rural (population density <600 per km^2^), peri-urban (population density 601 – 1400 per km^2^) and urban (population density >1400 per km^2^). All analyses were conducted using StataNow 19·5 MP-Parallel Edition (College Station, Texas, USA). This study is registered with ClinicalTrials.gov (NCT06017843).

## Ethical approval

Verbal informed consent was obtained from all participants at registration; study information was printed on the registration slip, read aloud, and participants provided verbal approval prior to screening. The institutional review boards of the Health Services Academy (No.7-82/IERC-HSA/2022-52), Pakistan and the Common Management Unit of Ministry of National Health Services, Regulations and Coordination (F. No 26-IRB-CMU-2023) approved the study.

## Results

Between August 2023 and September 2024, a total of 3,936 camps were conducted in the study districts of which 2,046 were in the control arm and 1,890 were in the intervention arm (Figure 2). Among camps conducted in the intervention arm, 1,095 (57.9%) were reported as being conducted with support of the AI software. Among these, 613 camps (56.0%) were verified as being conducted within a 5-km radius of the recommended locations and were included in the validated per-protocol analysis. Adherence to the intervention was not uniform across the van-teams and was highest in KPK (44.5%) and lowest in Baluchistan (8.3%) provinces (Supplementary Table 4). A higher proportion of camps were conducted in Punjab province in the intervention arm relative to the control (39.9% vs 26.0%) (Table 1). Community notable meetings were conducted more commonly in the intervention arm relative to the control (94·6% vs 68·7%, p-value <0·01).

**Table 1:** Baseline characteristics of camps.

|  | **Control**<br>**(N = 2,046)** | **Intervention**<br>**(N= 1,890)** | **p-value** |
| --- | --- | --- | --- |
| Province |  |  |  |
| Punjab | 532/2,046 (26.0%) | 755/1,890 (39.9%) | <0.01 |
| Baluchistan | 206/2,046 (10.1%) | 218/1,890 (11.5%) |  |
| KPK | 507/2,046 (24.8%) | 346/1,890 (18.3%) |  |
| Sindh | 801/2,046 (39.1%) | 571/1,890 (30.2%) |  |
| District Category** |  |  |  |
| Urban | 999/2,046 (48.8%) | 984/1,890 (52.1%) | 0.07 |
| Peri-urban | 510/2,046 (24.9%) | 463/1,890 (24.5%) |  |
| Rural | 537/2,046 (26.2%) | 443/1,890 (23.4%) |  |
| Community notables meeting |  |  |  |
| Conducted | 1,405/2,046 (68.7%) | 1,788/1,890 (94.6%) | <0.01 |
| Not conducted | 641/2,046 (31.3%) | 102/1,890 (5.4%) |  |

*\*\*Rural district defined as population density <600 per km<sup>2</sup>, peri-urban district define as population density of 601 – 1400 per km<sup>2</sup> and urban as population density >1400 per km<sup>2</sup>. Differences between groups assessed using chi-squared tests.*

**Figure 2:**
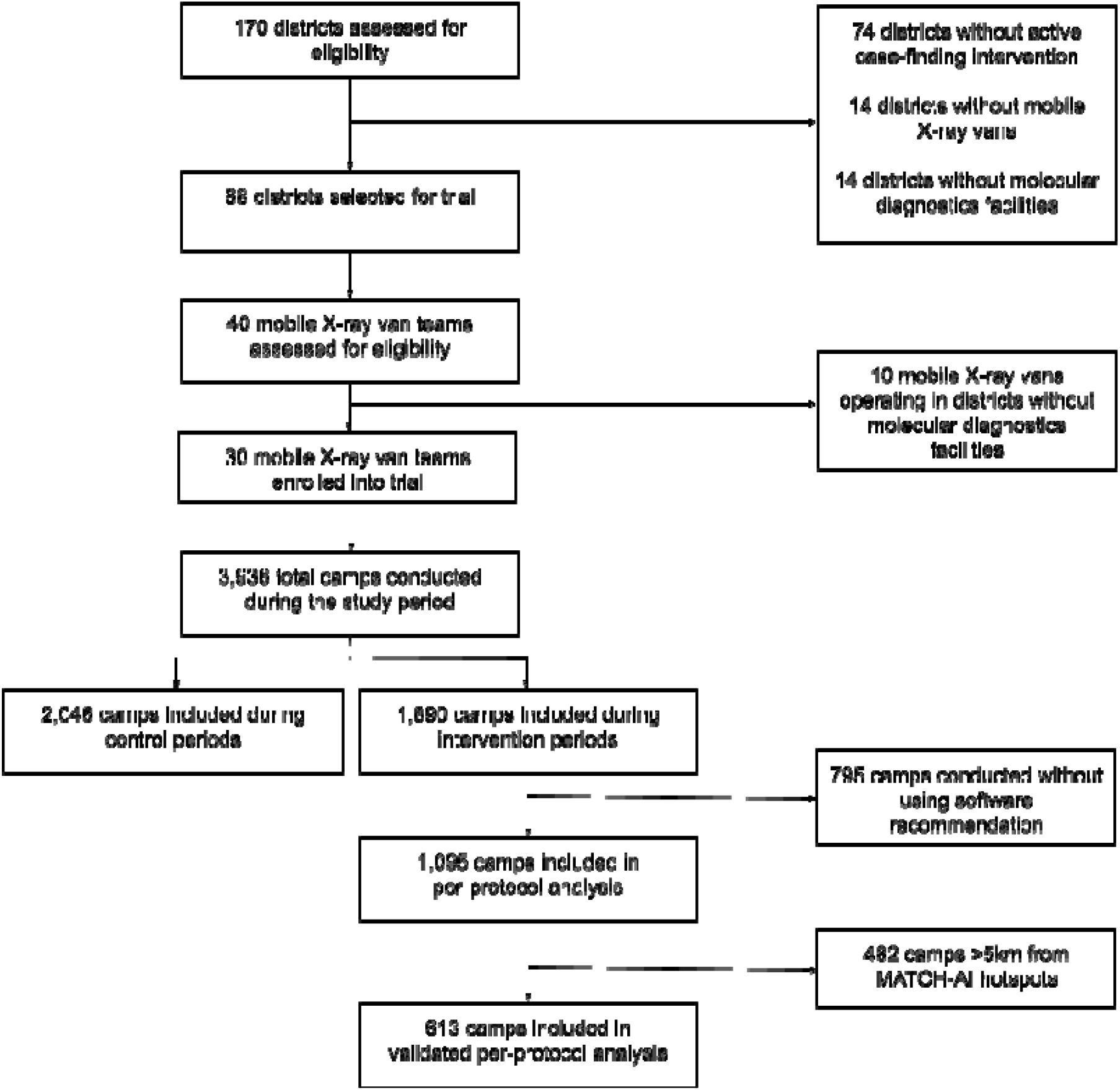
Trial profile.

**Figure 3:**
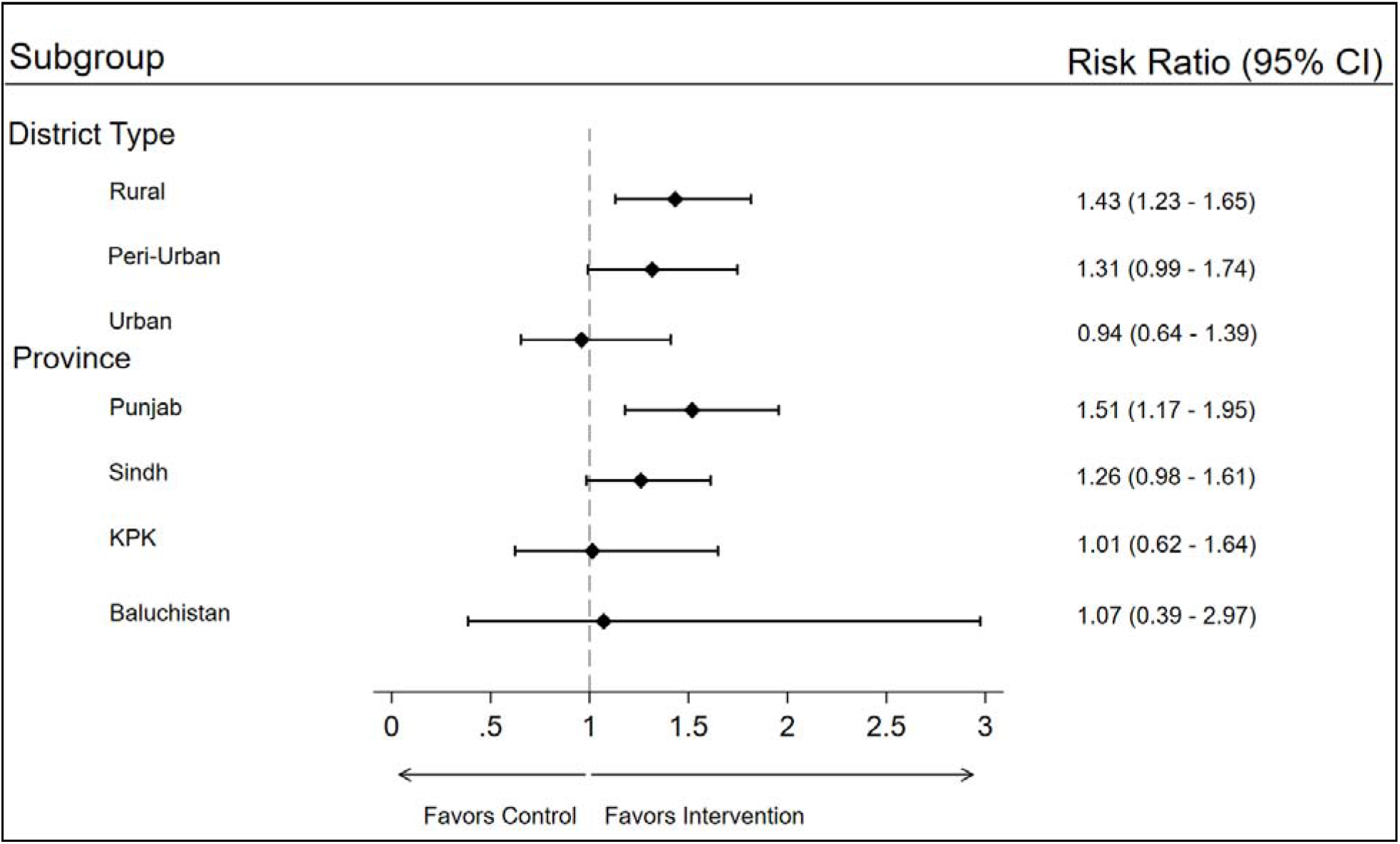
Post-hoc subgroup analyses for the primary outcome (validated per protocol) Risk ratios from adjusted analysis of camp positivity yield in the validated per protocol analysis. Rural districts defined as population density <600 per km^2^, peri-urban districts defined as population density of 601 – 1400 per km^2^ and urban districts defined as population density >1400 per km^2^.

A total of 269,254 individuals participated in the screening camps during the study among whom 142,359 (median per camp= 68, IQR: 55-80) were in the control arm and 126,895 (median per camp= 65, IQR: 54-78) were in the intervention arm (Table 2). Demographic characteristics of the participants were similar between the control and intervention arms. A total of 3,522 individuals with B+ TB were detected in the study of whom 1,923 (mean per camp= 0.94, SD= 1.20) were in the control arm and 1,599 (mean per camp = 0.85, SD= 1.11) were in the intervention arm. A total of 9,302 individuals with AF-TB were detected in the study of whom 5,053 (mean per camp= 2.47, SD= 1.83) were in the control arm and 4,249 (mean per camp = 2.25, SD= 1.88) were in the intervention arm. Detailed camp indicators and the care cascade are presented in the supplementary file.

**Table 2:** Baseline characteristics of participants.

|  | **Control**<br>**(N = 2,046)** | **Intervention**<br>**(N= 1,890)** | **p-value** |
| --- | --- | --- | --- |
| Total Participants, n (median per camp, IQR) | 142,359 (68, 55 – 80) | 126,895 (65, 54 – 78) | <0.01 |
| Participants' sex |  |  |  |
| Male | 70,265/142,359 (49.4%) | 60,958/126,895 (48.0%) | 0.08 |
| Female | 72,094/142,359 (50.6%) | 65,937/126,895 (52.0%) |  |
| Participants' age groups* |  |  |  |
| 0-15 years | 23,873/142,359 (16.8%) | 17,371/126,895 (13.7%) | <0.01 |
| 15 - 35 years | 45,933/142,359 (32.3%) | 41,197/126,895 (32.5%) | 0.12 |
| 35 - 50 years | 34,340/142,359 (24.1%) | 32,309/126,895 (25.5%) | 0.07 |
| >50 years | 38,213/142,359 (26.8%) | 36,018/126,895 (28.4%) | 0.53 |
| Individuals screened with CXR, n (median per camp, IQR) | 105,965 (50, 39 – 62) | 89,208 (47, 34 – 58) | <0.01 |
| Individuals tested with Xpert, n (median per camp, IQR) | 21,009 (9, 6 – 13) | 19,382 (9, 6 – 13) | 0.94 |
| Bacteriologically confirmed tuberculosis, n (mean per camp, SD) | 1,923 (0.94 ± 1.20) | 1,599 (0.85 ± 1.11) | 0.01 |
| All-Forms tuberculosis, n (mean per camp, SD) | 5,053 (2.47 ± 1.83) | 4,249 (2.25 ± 1.88) | <0.01 |

*\*Age was reported as a categorical variable in the aggregate camp dataset. Differences between groups assessed using chi-squared or t-tests.*

In the intention-to-treat analysis, the primary outcome, Camp Positivity Yield, was 7% higher in the intervention arm relative to the control (adjusted RR: 1.07, 95% CI: 0.94 – 1.22; p-value 0.27), however this difference was not statistically significant (Table 3). No significant differences or effect sizes were observed for any of the secondary outcomes in the ITT analysis. In the validated per-protocol analysis, Camp Positivity Yield was 32% higher in the intervention arm compared to the control (adjusted RR: 1.32, 95% CI: 1.12 – 1.54; p-value <0.01). Among secondary outcomes in the validated per-protocol analysis, Camp Positivity Rate was significantly higher in the intervention arm relative to the control (adjusted RR: 1.20, 95% CI: 1.02 – 1.41; p-value 0.03). A 7% benefit of the intervention was observed in the Camp All-Forms TB Yield, however, this difference was not statistically significant (adjusted RR: 1·07, 95% CI: 0·97 – 1·18; p-value 0.16).

**Table 3:** Primary and secondary outcomes.

|  | Unadjusted Analysis |  |  | Adjusted Analysis |  |  |
| --- | --- | --- | --- | --- | --- | --- |
|  | Risk / Rate Ratio | (95% CI) | p-value | Risk / Rate Ratio | (95% CI) | p-value |
| **Intention to treat analysis** |  |  |  |  |  |  |
| Camp positivity yield (Primary outcome) | 0.90 | (0.83 - 0.97) | 0.01 | 1.07 | (0.94 - 1.22) | 0.27 |
| Camp positivity rate | 1.00 | (0.92 - 1.08) | 0.94 | 1.01 | (0.89 - 1.15) | 0.84 |
| Camp All-Forms TB yield | 0.91 | (0.87 - 0.95) | <0.01 | 1.01 | (0.94 - 1.09) | 0.69 |
| Camp All-Forms TB rate | 1.00 | (0.96 - 1.04) | 0.96 | 0.94 | (0.88 - 1.02) | 0.12 |
| **Validated per protocol analysis** |  |  |  |  |  |  |
| Camp positivity yield (Primary outcome) | 1.22 | (1.10 - 1.36) | <0.01 | 1.32 | (1.12 - 1.54) | <0.01 |
| Camp positivity rate | 1.34 | (1.20 - 1.49) | <0.01 | 1.20 | (1.02 - 1.41) | 0.03 |
| Camp All-Forms TB yield | 1.01 | (0.95 - 1.07) | 0.75 | 1.07 | (0.97 - 1.18) | 0.16 |
| Camp All-Forms TB rate | 1.10 | (1.04 - 1.16) | <0.01 | 0.94 | (0.85 - 1.03) | 0.20 |

*Camp positivity yield is the number of individuals diagnosed with bacteriologically confirmed tuberculosis, per camp. Camp positivity rate is the number of individuals diagnosed with bacteriologically confirmed tuberculosis, per population screened with CXR.*
*Camp All-Forms TB yield is the number of individuals diagnosed with All-Forms of tuberculosis (including bacteriologically unconfirmed TB), per camp. Camp All-Forms rate is the number of individuals diagnosed with All-Forms of tuberculosis (including bacteriologically unconfirmed TB), per population screened with CXR.*
*Validated per protocol analysis is restricted to camps verified to be within 5-km radius of recommended locations by the software.*

**Table 4:** Pre-specified subgroup analysis for primary outcome.

|  | Unadjusted Analysis |  |  | Adjusted Analysis |  |  |
| --- | --- | --- | --- | --- | --- | --- |
|  | Risk Ratio | (95% CI) | p-value | Risk Ratio | (95% CI) | p-value |
| **Intention to treat analysis** |  |  |  |  |  |  |
| Low-yield districts | 0.92 | (0.77 - 1.09) | 0.34 | 0.97 | (0.74 - 1.28) | 0.84 |
| Medium-yield districts | 0.97 | (0.84 - 1.11) | 0.64 | 1.13 | (0.89 - 1.44) | 0.32 |
| High-yield districts | 0.76 | (0.68 - 0.85) | <0.01 | 0.97 | (0.80 - 1.17) | 0.71 |
| **Validated per protocol analysis** |  |  |  |  |  |  |
| Low-yield districts | 1.37 | (1.08 - 1.72) | 0.01 | 1.27 | (0.91 - 1.77) | 0.16 |
| Medium-yield districts | 1.41 | (1.15 - 1.72) | <0.01 | 1.57 | (1.13 - 2.18) | 0.01 |
| High-yield districts | 0.95 | (0.83 - 1.10) | 0.52 | 1.12 | (0.90 - 1.40) | 0.31 |

*Risk Ratios presented for camp positivity yield that refers to number of individuals diagnosed with bacteriologically confirmed tuberculosis, per camp. Districts with historical case-finding yields per population screened below 0.5% were classified as “Low-Yield”, those with 0.5% - 1% as “Medium-Yield” and those with >1% as “High-Yield.” Districts where screening data was not available were classified as Medium Yield.*

The pre-specified subgroup analysis is presented for the primary outcome. In the ITT analysis, Camp Positivity Yield was 13% higher in the intervention arm (adjusted RR 1.13, 95% CI: 0,89– 1.44; p-value 0.32) relative to the control in districts with historically medium yields, however, this difference was not statistically significant. No significant differences or effect sizes were observed for the remaining district categories in the ITT analysis. In validated per-protocol analysis, Camp Positivity Yield was 57% higher in the intervention arm (adjusted RR 1.57, 95% CI: 1.13–2.18; p-value 0.01) relative to the control. A benefit of the intervention was observed in low (adjusted RR 1.27, 95% CI: 0.91–1.77; p-value 0.16) and high (adjusted RR 1.12, 95% CI: 0.90–1.40; p-value 0.31) yield districts in the validated per-protocol analysis, however, these differences were not statistically significant.

Post-hoc analyses have been presented only for the adjusted validated per protocol analysis for the primary outcome (Figure 4). Yields in rural districts were 43% higher in the intervention arm relative to the control (adjusted RR 1.43, 95% CI: 1.23 – 1.65). Yields were 31% higher in peri-urban districts (adjusted RR 1.31, 95% CI: 0.99–1.74), however, this difference did not fully cross the superiority margin. In the provincial subgroup analysis, yields were significantly higher in Punjab (adjusted RR 1·51, 95% CI: 1·17 – 1·95). Yields were also higher in Sindh but this difference did not fully cross the superiority margin (adjusted RR 1·26, 95% CI: 0·98 – 1·61). No benefit was observed for the intervention in KPK and a small number of camps with verified GPS locations in Balochistan limited the precision of the intervention effect estimate.

## Discussion

This is the first prospective evaluation of a software designed to improve yields in community-wide systematic screening for TB through a geographical targeting approach. This highly pragmatic evaluation in routine, real-world implementation utilized a stepped wedge cluster randomized design, including 3,936 camps and involving 269,254 participants.

We found that providing AI-based targeting guidance for camp placement had nuanced results. The overall yield in intervention camps was not significantly different from control camps in the intention-to-treat analysis. This suggests that deviations from precise locations, i.e., beyond the recommended catchment radius of the predicted hotspots, substantially reduce yields, emphasizing the importance of precision during implementation. In the intention-to-treat analysis, which included all camps regardless of adherence to MATCH-AI recommendations, effect estimates were modest. This likely reflects that some intervention camps were conducted at locations that deviated from the AI-recommended sites, potentially diluting the overall impact.

However, when camps were conducted within 5 km of the targeting recommendations (per-protocol analysis), there was a 32% (adjusted RR: 1·32, 95% CI: 1·12–1·54) increase in adjusted yield. The larger effects observed in the validated per-protocol analysis therefore suggest that the effectiveness of AI-guided targeting depends on consistent adherence to recommended locations. These results underscore the importance of accurately targeting high-yield geographic locations in active case finding, and the importance of adherence to those targeting recommendations to maximize yield and (presumptively) cost-effectiveness of screening camps. This result supports the use of artificial intelligence such as MATCH-AI to guide screening efforts and improve diagnostic yields, when accompanied by efforts to support the implementation of targeting recommendations.

The divergence between intention-to-treat and per-protocol analyses illustrates how adherence to recommended sites influences outcomes and underscores that consistent implementation is key. While these findings explain what was observed, they do not explain why some targeting recommendations were implemented and others were not. Implementation was not affected by province, time, or specific screening van. Qualitative research has been conducted to understand the drivers of uptake of AI targeting recommendations and will be reported separately. Early findings suggest incomplete adherence to MATCH-AI recommendations due to operational constraints and limited understanding of the tool; existing partner commitments and low digital literacy further restricted optimal use, while field realities such as distance, poor access, repeated locations, and reliance on local knowledge were not fully captured by the AI model.

A previous retrospective analysis of the MATCH-AI software from Nigeria also demonstrated that model-based identification of high-risk communities increased TB yield, supporting the potential of data-informed targeting in ACF ^18^. This leads to two important conclusions. Firstly, it reinforces the importance of geographic targeting in ACF and during surveillance. This evidence aligns with available literature showing that systematic screening in geographically defined populations and pockets of undetected TB cases can maximize programmatic yield^19,20^. Tuberculosis is known to have a spatially heterogeneous distribution among communities, with cases clustering in specific geographic areas, and evidence suggests that a relatively small number of spots account for a disproportionate share of TB burden ^21^. Research suggests that untargeted screening may show minimal or no impact when case-finding efforts are not based on empirical evidence and are conducted in lower-risk areas ^4,22^. We have observed a similar situation in Pakistan in an ongoing analysis (unpublished), with a fairly large proportion of screening camps showing zero yield.

Secondly, our results add to the available evidence that the use of AI, which is essentially data-driven site selection by identifying locations with a higher probability of TB, can positively impact case-finding efforts in high-burden settings ^19^. MATCH-AI, in comparison to the usual site selection approach, systematically integrates multiple data sources such as prior TB notifications, key socio-demographic and geographic indicators, and spatial correlations, and presents an unbiased identification of high-risk locations that might be overlooked by human judgment. Human site selection may rely on information influenced by convenience, political considerations, or familiarity with certain communities, potentially leading to repeated screening in the same locations with diminishing returns. Together, these conclusions emphasize that integrating spatial intelligence into routine active case finding is not merely an optimization but may be a necessary component of effective tuberculosis surveillance and elimination.

The subgroup analysis suggests that the software performed better in districts with moderate pre-existing yields of TB, while it was less effective in districts where yields were already high or low. In high-yield areas, since the burden of TB is already high, any reasonable screening strategy should capture a large proportion of cases, while in districts with low yield the underlying TB prevalence may be too low to identify additional cases. Alternatively, staff may already have been familiar with potential spots in low-yield districts, limiting the incremental benefit from the AI tool. The potential benefit of MATCH-AI in medium-yield districts is an important finding, as these districts require greater precision during ACF and represent settings where the potential for TB case detection exists but is not fully realized by routine selection approaches. Provincial analyses indicated higher performance in Punjab and Sindh, as well as in rural districts, suggesting identification of previously unknown spots of TB by the software. Variability in camp outcomes and participation is also influenced by operational factors including camp timing, camp location, and pre-camp awareness-raising activities, which can affect positivity rates independently of site selection, as noted in other active case-finding studies. Because AI-recommended sites were frequently new to field teams and staff targets are based on attendance numbers, additional mobilization efforts such as additional area notable meetings may have been undertaken in the intervention arm.

It is important to note that the control arm in this study also applied a targeting strategy, drawing on facility-level notification data as well as the prior knowledge and experience of healthcare workers in Pakistan. These teams have several years of experience in conducting ACF and may have been familiar with potential TB hotspots in their districts. Despite relatively high baseline ACF yields (0·88 per camp), the intervention demonstrated measurable improvements in camps adhering to AI-recommended locations. However, notification data alone is not a reliable indicator, as it is heavily influenced by the capacity and quality of diagnostic and treatment services in a given area. Regions with weaker services may report fewer cases, which does not necessarily reflect lower TB transmission.

### Strengths

Some of the key strengths of this study include the application of AI within pragmatic real-world deployment supported by a large sample size of over 250,000 individuals screened. Conducted under routine operational conditions in a high TB incidence country such as Pakistan, the study provides evidence that is highly relevant and generalizable to similar high-burden contexts. All cases identified during the study were reported to the NTP, and this synergistic approach allowed greater ownership of the intervention. In addition, the trial supported capacity-building for implementation science research in Pakistan.

### Limitations

This trial had several limitations. Due to the scale of the ACF program, we did not conduct independent monitoring or quality assurance audits of laboratory testing for microbiological outcomes. However, laboratory technicians involved in ACF receive regular training on Xpert testing, which is reviewed by the NTP. A review of individuals initiated on treatment empirically with bacteriologically unconfirmed TB was not conducted, and this may have affected the results of the all-forms TB outcomes that did not show significant benefit. This trial further demonstrates the need for standardization of management of bacteriologically unconfirmed TB in programmatic settings.

It is important to note that while this trial evaluated the use of a targeted approach toward ACF, it relied on a particular software to guide decision-making, and it is likely that alternative platforms or prediction models would lead to different location recommendations.

## Conclusion

This study demonstrated that geographical targeting using AI-based software improved yields of bacteriologically confirmed TB detected during active case finding in the validated per-protocol analysis. Importantly, while AI offers considerable potential to improve the efficiency and yield of TB active case finding, its effectiveness can be enhanced by integration with contextual human knowledge and local experience.

Our results support existing evidence that the distribution of TB is spatially heterogeneous and that programs should consider geographical targeting of their interventions. The study utilized a readily available commercial software that may be suitable in similar settings for other high- and medium-burden countries, particularly in settings with limited prior ACF experience.

## Acknowledgements

This work was supported, in part, by the Gates Foundation INV-037454. The conclusions and opinions expressed in this work are those of the authors alone and shall not be attributed to the Foundation. Under the grant conditions of the Foundation, a Creative Commons Attribution 4.0 License has already been assigned to the Author Accepted Manuscript version that might arise from this submission. Please note works submitted as a preprint have not undergone a peer review process.

## Author Contributions

AL and FE were involved in the study conception. AM, SMAZ, FE, TR were involved in study design and protocol development. AL, AT, SN and NN were responsible for study implementation and oversight. SMAZ, FR and WA were the study statisticians. SMAZ developed the statistical analysis plan and prepared the results. AM, WA, AS and NN accessed and verified the data and were involved in the analysis. FA and NN were involved in data monitoring. AL and AT were involved in project management. AL and FE were involved in scientific coordination. FE, SMAZ, AM and WA were involved in interpreting the results. AM and SMAZ wrote the first draft. All authors provided comments, read and approved the manuscript. All authors had full access to all the data in the study and had final responsibility for the decision to submit for publication.

## Data availability

All data supporting these findings can be found online at Figshare https://doi.org/10.6084/m9.figshare.32333712 and https://github.com/smazaidi87/SPOT-TB-Trial-Analysis-Code.git. Data are available under the terms of the Creative Commons Attribution 4.0 International License (CC BY 4.0).

## Use of Generative AI

The authors used generative AI and AI-assisted technologies only for language editing to improve readability during manuscript preparation. The authors reviewed and edited the content as needed and take full responsibility for the content of the publication.

